# Improving postsurgical fall detection for older Americans using LLM-driven analysis of clinical narratives

**DOI:** 10.1101/2024.06.25.24309480

**Authors:** Malvika Pillai, Terri L Blumke, Joachim Studnia, Yuqing Wang, Zachary P Veigulis, Anna D Ware, Peter J Hoover, Ian R Carroll, Keith Humphreys, Thomas F Osborne, Steven M. Asch, Tina Hernandez-Boussard, Catherine M Curtin

## Abstract

Postsurgical falls have significant patient and societal implications but remain challenging to identify and track. Detecting postsurgical falls is crucial to improve patient care for older adults and reduce healthcare costs. Large language models (LLMs) offer a promising solution for reliable and automated fall detection using unstructured data in clinical notes. We tested several LLM prompting approaches to postsurgical fall detection in two different healthcare systems with three open-source LLMs. The Mixtral-8×7B zero-shot had the best performance at Stanford Health Care (PPV = 0.81, recall = 0.67) and the Veterans Health Administration (PPV = 0.93, recall = 0.94). These results demonstrate that LLMs can detect falls with little to no guidance and lay groundwork for applications of LLMs in fall prediction and prevention across many different settings.

## INTRODUCTION

Falls are a leading cause of morbidity and mortality in all age groups, but disproportionately in older adults^1,2^. Fall-related injuries in older adults are increasing and have numerous negative consequences to the patient and enormous social costs^3^. In 2015, falls cost the United States healthcare system $50 billion per year, and that cost is expected to double by 2030 with several expert bodies calling for fall prevention protocols^4-6^. As a result, understanding and preventing falls is a high priority at every level.

Due to the lingering effects of anesthesia medications and associated physiologic changes of surgery, the postoperative patient is at a disproportionate increased risk for falls. Postsurgical falls are a tempting target for prevention efforts. They are common (1-4% of patients) and can result in morbidity and even surgical failure. The discrete timing of surgery allows easy identification of the risk period initiation, easing implementation of targeted fall reduction strategies.

Before such strategies can be implemented, it is critical to understand who and how patients fall. Traditionally, studying clinical conditions for large numbers of patients requires assessing the structured data of specific medical diagnostic codes in the electronic health record (EHR). However, falls are often recorded in the unstructured free text of the written narrative, making it difficult to assess. Although for some specific coded injuries, such as a wrist fracture that is typically associated with a fall, inferences can be drawn that suggest a fall is a likely cause. However, when there is not a specific injury coded, the incidence of falls becomes much more opaque. Therefore, advanced analytics methods that can extract insights from unstructured data are needed to understand the true incidence, risks, and therefore optimal prevention strategies for falls.

In this study, we aim to identify fall events in clinical notes using open-source large language models (LLMs) that can interact with the free-text electronic medical records across healthcare systems. To this end, we developed an approach that utilizes LLMs for binary classification to detect postsurgical falls. We hypothesize that these LLMs will accurately detect and classify fall events, deepening our understanding of who is at risk and improving our ability to predict and prevent falls in the future.

## METHODS

### Setting and data collection

This retrospective cohort study was performed at a large academic healthcare system. The study was approved by the institutional review board at Stanford University (Stanford University, Stanford, CA, USA; IRB-34551) and follows the Minimum Information for Medical AI Reporting (MINIMAR) guidelines^7^. This study was validated at the Veterans Health Administration (VHA), the largest integrated healthcare system in the United States with 172 medical centers across the country.

This study included EHR data from patients who underwent a surgical procedure between 2012-2022 at Stanford Health Care and between 2020-2022 at the VHA (Figure 1). The patients included were at least 50 years of age with an inpatient stay of 90 days or less with no death reported within 30 days after surgery (n = 59,047). All patient notes dated after surgery and within the observation period (e.g., progress notes, nursing notes) were utilized in this analysis. For patients who underwent more than one surgery within the observation period, only the first surgery was extracted. For external validation in the VHA, data were extracted for patients who underwent a surgical procedure between 2020-2022 who met the inclusion criteria described above (n=135,966).

**Figure 1.**
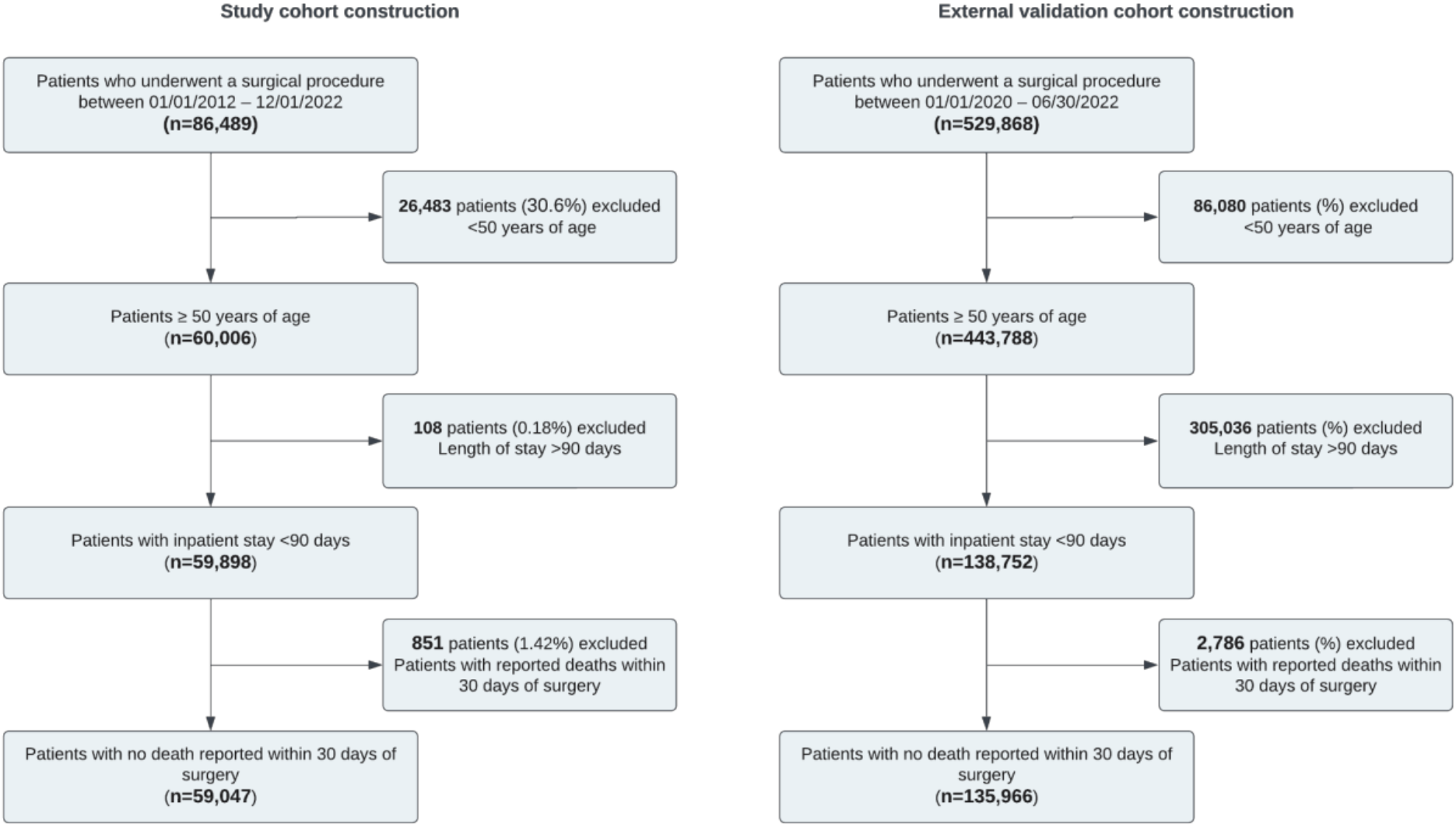
Cohort flowchart detailing study participants included in the fall detection analysis

### Study cohorts

The internal study cohort included 59,047 patients who underwent 75,179 surgeries at a large academic medical center (Figure 1, Table 1). For our defined age group (≥ 50 years of age), the median age at surgery was 66 years, and 52% of patients were female. Non-Hispanic White (63%), Non-Hispanic Asian (13%), and Hispanic (12%) patients comprised the majority of the cohort. At the time of surgery, 59% of patients were privately insured. The external validation cohort included 135,966 patients who underwent 138,752 surgeries and had 189,442 associated encounters at VHA (Figure 1, Table 1). The median age at surgery was 70 years, and 94% of patients were male. Non-Hispanic White (68%) and Non-Hispanic Black (20%) patients comprised the majority of the cohort. Patient characteristics from both cohorts are described in detail in Table 1.

**Table 1.**
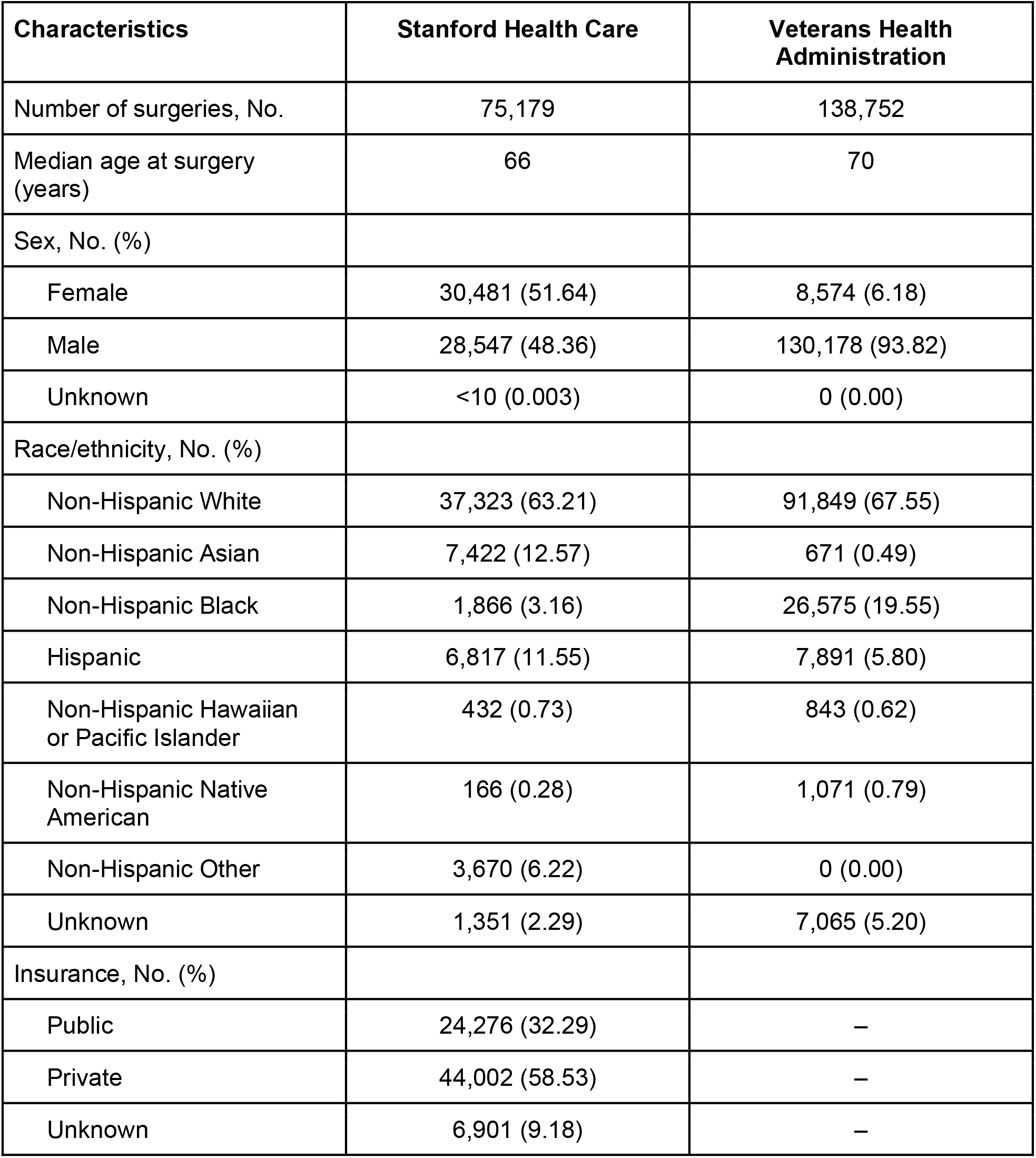
Patient cohort characteristics.

### Data preparation

We processed the notes through a rule-based approach to create the dataset for annotation. We used ICD-9-CM and ICD-10-CM diagnosis codes to identify falls in structured data and used regular expressions (regex) to identify fall mentions in clinical notes. Patterson et al (2019)^8^ defined regex to identify and categorize fall mentions, and we iteratively updated the published regex with terms from Stanford data to make it more robust. We used the regex protocol as a preliminary note filter. To define a sample for annotation, we took a sample of notes with regex-identified falls, regex-identified and code-identified falls, and no fall mentions at random. The regex protocol is described in Appendix 1.

### Manual chart review and annotation

To create a gold standard dataset to validate our approach, we developed standardized annotation guidelines and two healthcare professionals conducted manual chart review and annotations. Notes were annotated (human-labeled) as containing a fall event, history of falls, fall risk, other mentions of the word “fall”, or no fall. In the annotation guidelines, we specified granular sub-categories for negative cases to enable a thorough error analysis to understand model performance (Table 2).

**Table 2.**
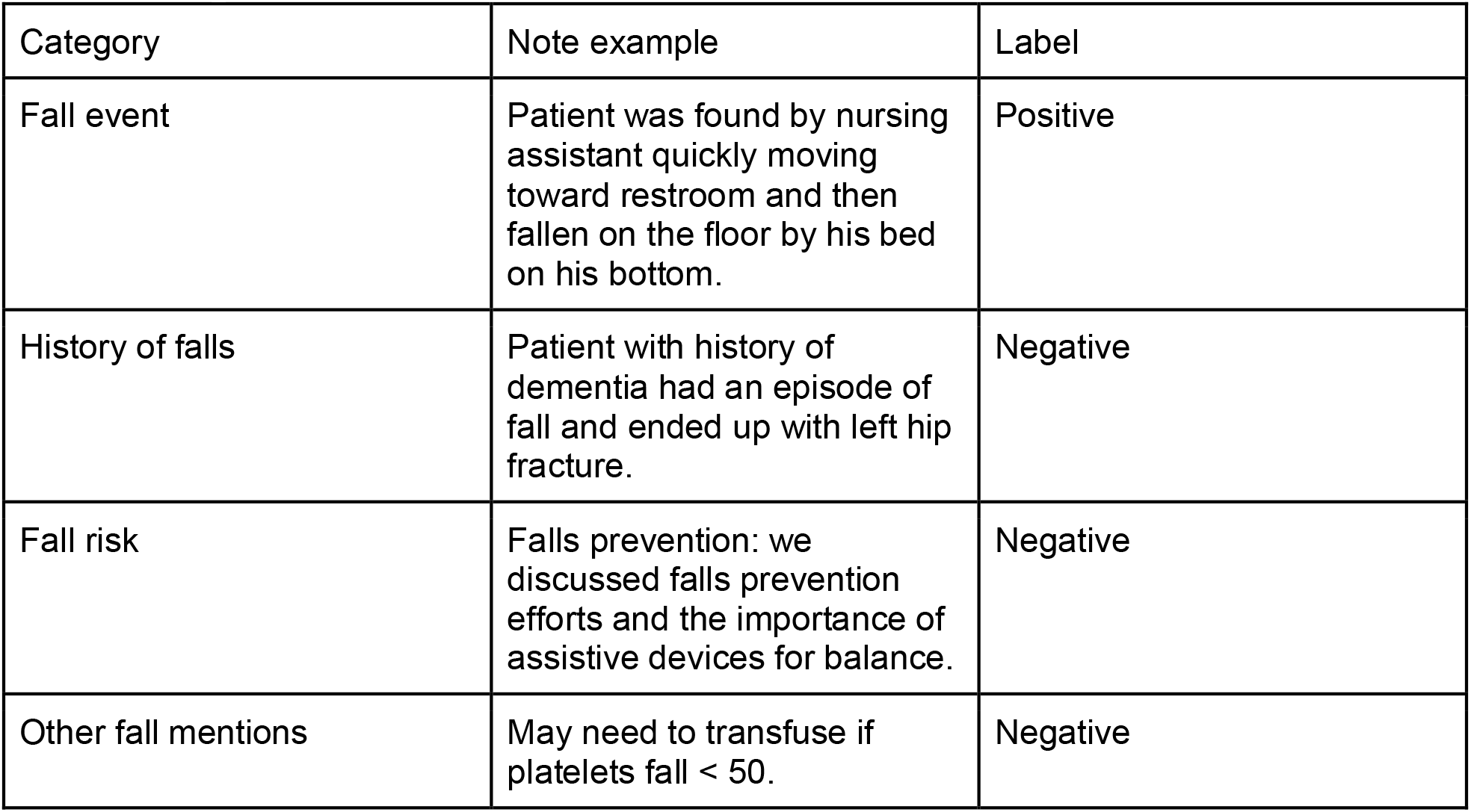
Categories defined for annotation.

| Category | Note example | Label |
| --- | --- | --- |
| Fall event | Patient was found by nursing assistant quickly moving toward restroom and then fallen on the floor by his bed on his bottom. | Positive |
| History of falls | Patient with history of dementia had an episode of fall and ended up with left hip fracture. | Negative |
| Fall risk | Falls prevention: we discussed falls prevention efforts and the importance of assistive devices for balance. | Negative |
| Other fall mentions | May need to transfuse if platelets fall < 50. | Negative |

As the task in this study was to identify fall events, our main objective was to distinguish fall events from other types of fall mentions. The sentence before, the annotated sentence, and the sentence after were extracted for each regex-tagged fall as a note chunk. If a note contained more than one fall mention, all note chunks were concatenated. For classification, the annotated categories were grouped into two outcome classes: fall event and no fall event. The fall event category consisted of patient falls. The no fall event category consisted of history of falls, fall risk, other mentions, and no fall. Cohen’s kappa was selected to measure inter-annotator agreement (IAA).

### Models

We utilized three state-of-the-art LLMs: Large Language Model Meta AI (LLaMA) 3-8B^9^, Gemma-7B^10^, and Mixtral-8×7B^11^. All models selected for this analysis were run in two settings on HIPAA-compliant machines locally with the internal study cohort (large, academic healthcare system) and through cloud computing with the external validation cohort (national, integrated healthcare system).

### Fall detection approaches

To enable large-scale note analysis without heavy computation and avoid annotation burden, we used two prompting approaches to test the capability of LLMs for binary fall detection: zero-shot prompting (i.e., asking the model questions without any further tuning) and few-shot prompting (i.e., in-context learning by providing the model with a small sample of annotated notes before asking questions). The zero-shot prompt was developed with an iterative process based on the annotation guidelines described above. The final prompts were created by examining response accuracy and completeness after each prompt iteration. We then provided a small amount of labeled data for few-shot prompting and assessed performance.

Experimental pipelines were created using the internal study cohort and replicated in the external validation cohort. From each cohort, we incorporated three example notes in the prompt, and both sets of three followed the same format. Two notes contained different types of fall events and one note did not contain a fall event. The final prompts are described in Appendix 2. Using a data post-processing pipeline, we cleaned the responses from each model and extracted the binary outputs.

### Internal validation (Academic Health System)

The models were deployed on inpatient notes from all patients in the cohort (n=59047 patients, 8,394,866 clinical notes). The notes were processed through the regex protocol to create shortened notes as previously described. Sentences with regex-tagged falls and the sentences before and after them were taken as note chunks. For notes with multiple fall mentions, all note chunks were concatenated. To evaluate model performance, we randomly selected a sample of clinical notes for annotation, creating a gold standard dataset. We organized brainstorming sessions with clinicians and data scientists to define falls and develop annotation guidelines.

Using an iterative process, we refined the guidelines and achieved consensus among annotators. Initially, two healthcare professionals independently annotated 50 notes using the guidelines. All disagreements were reviewed and discussed among the annotators, leading to guideline revisions. Subsequently, another 50 notes were independently annotated, and IAA was evaluated over 100 notes (IAA=0.84). Once consensus was reached, an additional 500 notes were annotated, resulting in a gold standard dataset of 600 notes. Following metric-based performance evaluation, we conducted an error analysis of common misclassifications across models to assess performance.

### External validation (VHA)

Models were validated on a sample of 189,442 encounters with 25,785,498 clinical notes from VHA between 2020-2022. To efficiently evaluate our approach, models were run on a targeted sample of notes. We defined three categories for targeted sampling: likely positive, possibly positive, and likely negative notes. VHA sites use official fall note templates to report incidences of falls. We first separated patients into two groups: those having an official fall report (5,516 encounters) or not (183,926 encounters). We defined notes within 24 hours of an official fall report as likely positive and notes within one month prior to an official fall report as possibly positive. Since these notes were written around the time of an official fall report, we hypothesized that the falls physicians or nurses would write about in surrounding notes would be related to postsurgical falls, rather than other types of fall mentions. Notes from patients without official fall reports were defined as likely negative. Following the gold standard annotation guidelines, a sample of 200 notes was manually reviewed by two healthcare professionals, and IAA was evaluated for a subset of 50 notes (IAA=0.95).

## RESULTS

Models were evaluated using precision (i.e., positive predictive value (PPV)), recall (i.e., sensitivity), and area under the receiver operating characteristic curve (AUROC).

### Internal validation results

#### Regular expression protocol

All patient notes from the internal study cohort were passed through the regex protocol. Of the 59,047 patients in the cohort, 14,550 (25%) patients had at least one note that met the regex criteria, which is much higher than the number of patients with an ICD-CM code for falls (2,345 patients). Of the 2,345 patients with a fall code, 1,812 patients (77%) had at least one note that met the regex criteria. A subset of notes for patients with a fall code but without a fall mentioned in the notes were manually examined. Common reasons for not having a fall mention within the note but having an ICD-CM fall code were: 1) poor fall reporting such that no fall was mentioned in the note, and 2) mentions of fall precautions, which were excluded by regex.

#### Binary classification results

Models were deployed on all regex-flagged notes in the internal study cohort (n=321,685), and the gold standard dataset (600 randomly selected notes from 348 patients) was used to evaluate the models. Of the 600 notes, 56.7% (n=340) notes were labeled as not containing a fall event. Between zero and few-shot approaches, the models achieved higher performance when provided few-shot prompts than when provided zero-shot prompts. Mixtral-8×7B few-shot had the highest precision, and Gemma-7B few-shot had the highest recall. Gemma-7B and LLaMA3-8B few-shot had the highest AUROC (Table 3). However, in terms of balanced performance while favoring precision over recall, we found that Mixtral-8×7B zero-shot was the best model.

**Table 3.** Internal validation at Stanford Health Care: fall classification performance.

| Few-shot prompting results |  |  |  |
| --- | --- | --- | --- |
| Model | Precision | Recall | AUROC |
| LLaMA3-8B | 0.72 | 0.90 | 0.82 |
| Gemma-7B | 0.71 | 0.93 | 0.82 |
| Mixtral-8x7B | 0.92 | 0.56 | 0.76 |
| Zero-shot prompting results |  |  |  |
| LLaMA3-8B | 0.78 | 0.70 | 0.77 |
| Gemma-7B | 0.71 | 0.87 | 0.79 |
| Mixtral-8x7B | 0.81 | 0.67 | 0.78 |

#### Error analysis

Common misclassifications across models were examined to understand potential failure cases. From both prompting approaches, false positive classifications were mostly due to historical fall mentions. Of the 20 notes all models misclassified as positive from zero-shot prompting, 85% (n=17) contained historical fall mentions, 10% (n=2) contained miscellaneous fall mentions, and 5% (n=1) contained fall risk mentions. Using few-shot prompting, all models misclassified 8 notes which contained historical fall mentions.

Table 4 describes misclassification examples from both zero-shot and few-shot prompting results, categorized by major error type. Our primary goal was to maximize precision while maintaining a balance with recall, so various false positive cases are illustrated. The model explanations provide insight into the reasoning behind the model’s output and are presented in the table. In the historical fall examples, note H1 was incorrectly labeled as positive with a zero-shot approach; with a few-shot approach, note H2 was incorrectly labeled as containing falls, and note H3 contains a historical mention of a fall that Mixtral-8×7B correctly labeled but other models incorrectly labeled. In the miscellaneous fall mention example, note M1 contains a note where the model failed to disambiguate another use of the word “fell” and incorrectly labeled the note as containing a fall mention. We indicate the possible reason for misclassification in **bold** in the note.

**Table 4.** Example notes from false positive classifications from zero-shot and few-shot prompting experiments grouped by error type (edited to prevent reidentification).

| # | Note | Explanation | Output | True Label |
| --- | --- | --- | --- | --- |
| **Historical fall** |  |  |  |  |
| H1 | Hospital admissions for DKA or severe hypoglycemia: **early YYYY fell and bruised his left temple as well as broke front tooth...** History of scheduled stress test was deferred given he was in tachycardia and went to emergency room for evaluation... Has upcoming trip to Niagara Falls on MM/DD. Reports currently taking cholesterol medication, and tolerating well. | <u>**Zero-shot**</u><br>Gemma-7B:<br>The patient had a fall in early YYYY, resulting in a bruised left temple and a broken front tooth.<br>LLaMA3-8B:<br>The patient had a fall event mentioned in the note, specifically "fell and bruised his left temple as well as broke front tooth" during an admission for DKA or severe hypoglycemia early in YYYY.<br>Mixtral-8x7B:<br>The note mentions a fall event that occurred in early YYYY when the patient fell and bruised his left temple as well as broke his front tooth. | Gemma-7B: 1<br>LLaMA3-8B: 1<br>Mixtral-8x7B: 1 | 0 |
| H2 | Patient also reported to this nurse he has had **frequent falls at home**. One recently, causing the wife to fall as well while assisting him. Concerned with patient's mobility as he seems unsteady while ambulating to | <u>**Few-shot**</u><br>Gemma-7B:<br>The patient has had frequent falls at home, including one that caused his wife to fall.<br>LLaMA3-8B:<br>The patient reported having frequent | Gemma-7B: 1<br>LLaMA3-8B: 1<br>Mixtral-8x7B: 1 | 0 |
|  | bathroom. Will report these findings to MD. | falls at home, including one recent incident where his wife fell while assisting him.<br>Mixtral-8x7B:<br>The patient reported frequent falls at home, one of which occurred recently. |  |  |
| H3 | CHIEF COMPLAINT: A gentleman presents for followup of low back pain and left lower extremity pain. HISTORY OF PRESENTING ILLNESS: Mr. X is a very pleasant X-year-old gentleman, well known to the clinic for approximately 2 years of progressive and worsening low back pain and left lower extremity pain. **His history, in brief, his pain started after a fall.** He has undergone 4 transforaminal epidural steroid injections. | **Few-shot**<br>Gemma-7B:<br>The patient had a fall that led to low back pain and left lower extremity pain.<br>LLaMA3-8B:<br>The patient's low back pain and left lower extremity pain started after a fall.<br>Mixtral-8x7B:<br>The note mentions that the patient's pain started after a fall, but it does not specify whether the patient fell after surgery or provide any details about a fall event after surgery. | Gemma: 1<br>LLaMA3-8B: 1<br>Mixtral-8x7B: 0 | 0 |
| **Miscellaneous fall mention** |  |  |  |  |
| M1 | Patient says "I am doing OK." He was started on a diet, which he is tolerating. **Ate a quiche, but then fell full after 5 bites** and stopped. Denies nausea. Does have some painful bloating. | **Zero-shot**<br>Gemma-7B:<br>The patient fell after eating 5 bites of quiche.<br>LLaMA3-8B:<br>The patient had a fall event mentioned in the note, specifically "fell full after 5 bites and stopped".<br>Mixtral-8x7B:<br>The patient fell while eating quiche, as indicated by "he then fell full after 5 bites and stopped." This is a fall event. | Gemma-7B: 1<br>LLaMA3-8B: 1<br>Mixtral-8x7B: 1 | 0 |

### External validation results (VHA)

#### Regular expression protocol

There were 30,766 notes within 24 hours after a designated fall note (likely positive) from 2,205 patients, and 7,861 notes passed the regular expression protocol. 2,346 patients had 392,511 notes within one month prior to a designated fall note (possibly positive), and 55,997 notes passed the regular expression protocol. 134,373 notes were selected from 133,557 patients who did not have a designated fall note (likely negative), and 11,040 notes passed the regular expression protocol. Notes were randomly selected from each targeted sampling category (likely positive, possibly positive, likely negative) to create the annotated dataset (n=200). Of these 200 randomly selected notes, 80 notes were labeled as not containing a fall event and 120 notes were labeled as containing a fall event to construct the gold standard validation set.

#### Model classification results

Model performance was evaluated across the gold standard validation set over 5 model runs. Model performance was identical across 5 runs for all models. Between zero-shot and few-shot prompting approaches, using zero-shot prompting achieves higher balanced performance while still favoring precision (Table 5). Mixtral-8×7B few-shot had the highest precision, LLaMA3-8B had the highest recall, and Mixtral-8×7B zero-shot had the highest AUROC. In terms of balanced results, Mixtral-8×7B zero-shot had the best performance.

**Table 5.** External validation at VHA: fall classification performance.

| Few-shot prompting results |  |  |  |
| --- | --- | --- | --- |
| Model | Precision | Recall | AUROC |
| LLaMA3-8B | 0.98 | 0.70 | 0.84 |
| Gemma-7B | 0.62 | 0.83 | 0.54 |
| Mixtral-8x7B | 0.99 | 0.57 | 0.78 |
| Zero-shot prompting results |  |  |  |
| LLaMA3-8B | 0.88 | 0.99 | 0.90 |
| Gemma-7B | 0.94 | 0.84 | 0.88 |
| Mixtral-8x7B | 0.93 | 0.94 | 0.92 |

## DISCUSSION

This retrospective national observational study used state of the art generative artificial intelligence methods to streamline postsurgical fall identification efforts across healthcare systems. We found that the LLMs were able to identify postsurgical falls with precision, recall and AUROCs that are comparable to manual chart review in many settings^8^. To establish a pipeline and investigate the potential of LLMs to aid in postsurgical fall detection, we conducted experiments that tested different prompting techniques. Tailoring prompting approaches across health care systems with distinct note formats and cultures improved performance. Few-shot prompting outperformed zero-shot (“out of the box”) prompting in the academic center, while zero-shot prompting significantly surpassed few-shot prompting in VHA.

We had expected few-shot prompting to achieve higher performance across settings. One possible explanation is that the VHA dataset contains notes from multiple sites with site-specific note templates and standardization, but only three example notes were used for few-shot prompting. To improve performance with few-shot prompting, the models may need examples of diverse notes across sites to learn better.

Zero shot prompting may, in any case, be a more generalizable application of LLMs to fall detection. Model performance varies greatly as a result of minute changes to input prompts, making generalizability a concern when crafting institution-specific or broadly applicable prompts. Specifically, when incorporating few-shot examples into prompts, the model may produce outputs that mimic the example styles rather than more faithfully recognizing patterns in the input data. This could introduce bias by causing the model to favor some patterns over others. If future research produces comparable performance from external validation at scale, it would serve as proof of concept for the transferability of our approach.

LLMs are powerful and can be used for fall detection. Current literature has explored other tools that have identified fall events within controlled settings with very high sensitivities and specificities^12,13^. However, these approaches have largely been institution-specific and have relied on large annotated datasets which are cumbersome to produce. Cheligeer et al. (2024)^14^ developed a Bidirectional Encoder Representations from Transformers (BERT)-based approach for inpatient fall detection, producing three models optimized for high sensitivity (97.7% sensitivity, 15.6% PPV), high PPV (27.9% sensitivity, 85.7% PPV), and high F1-score (66.7% sensitivity, 60.5% PPV). Their approach examined how to balance false positives and false negatives with multiple models. Kawazoe et al. (2022)^12^ developed an approach using structured data and BERT to classify clinical notes written in Japanese to detect and predict falls. Their approach had good sensitivity (0.74) and specificity (0.84) but low PPV (0.09).

Patterson et al. (2019)^8^ developed a high-performing regular expression-based approach at a single site, with a focus on being easy to implement and adapt (95.8% sensitivity, 92.0% PPV). Building on this work, LLMs represent a tool that can effectively detect time bound falls without tremendous manual chart review in a generalizable way.

The fall identification work is an important first step to predicting postoperative fall risk^15^. When we can understand who falls at a population level, we can build important clinical prediction models that identify and incorporate various fall risks, so we can optimally tailor our prevention interventions^16,17^. For instance, a patient with a high fall risk may benefit from a wearable device that monitors their movements and alerts nursing staff to any unusual activity, while a patient with a low fall risk might only require non-slip socks. Past efforts to identify falls in clinical narratives have relied on vast amounts of clinician annotated notes to train natural language processing models. We present an approach that alleviates the burden on subject matter experts without sacrificing model performance and research quality, which could reduce cognitive overload in gauging ground truth. Additionally, due to local differences in documentation, previously documented identification methods have shown limited generalizability. We leveraged prompting approaches with a small, annotated set and demonstrate generalizability across two different healthcare settings. By utilizing LLMs, we can enhance fall detection to predict and prevent falls. This not only streamlines the workload for healthcare professionals but also ensures that fall incidents are accurately identified, allowing for effective interventions based on individual risk profiles. Our approach paves the way for broader application and integration of advanced models in diverse clinical settings, ultimately contributing to improved patient safety and care outcomes.

### Limitations

This study has several limitations. First, while LLMs are highly generalizable due to their size, their performance may be constrained by site-specific data. For instance, the findings from the VHA might not be applicable to other populations due to differences in note styles compared to settings like academic health systems. Second, we did not engage in extensive prompt engineering to further optimize performance. Additional iterations of prompt engineering and the use of other strategies, such as chain-of-thought prompting, could potentially enhance model performance. Third, the study was conducted within a specific time frame, and changes in clinical practices or data recording methods over time could affect the results. Future research should include periodic model re-evaluation to ensure its relevance over time. Fourth, there are inherent limitations related to the black-box nature of LLMs and potential biases in the text they were trained on, which could have unforeseen consequences in our analysis. Given the challenges in assessing model bias and fairness, it is crucial for healthcare organizations aiming to implement LLM-based systems to recognize and mitigate these potential pitfalls.

### Future directions

In this study, we present an approach to identify postsurgical falls in clinical narratives. This is intended to lead to future work that includes predicting fall risk and identifying key predictors of falls. We aim to develop a pipeline that can be deployed across healthcare sites for continuous risk evaluation and re-evaluation based on changes in patient conditions, setting risk thresholds that can be used for fall prevention strategy selection. In future work, we also plan to assess perceptions of employing data-driven tools for fall risk assessment compared to using manual forms of assessment.

## Data Availability

Due to US Department of Veterans Affairs (VA) regulations and ethics agreements, the data utilized for this assessment are not permitted to leave the VA firewall without a Data Use Agreement. However, VA data can be made available to researchers with an approved IRB and VA authorized study protocol. For more information, please visit https://www.virec.research.va.gov or contact the VA Information Resource Center at

## Acknowledgements

We would like to acknowledge Deborah Kenney for her invaluable assistance with the annotation process.

## Author Contributions

MP, TH-B, and CMC contributed to the conceptualization of the study. MP, TH-B, and CMC contributed to study design. JS and TLB contributed to data curation. MP, JS, and YW contributed to data analysis. MP, TLB, ZPV, ADW, PJH, TFO, SMA and CMC contributed to validation of the study. CMC, IC, KH, TFO, and SMA provided the clinical expertise. MP drafted the original manuscript, and all authors contributed to critical review of the manuscript.

