## Appendices for "Improving postsurgical fall detection for older Americans using LLM-driven analysis of clinical narratives"

### Appendix 1. Regular expression protocol

| **Category** | **Example** |
| --- | --- |
| Original inclusion regex | Sure fall:  '(before\|after\|prior to\|while) (the )?fall(ing)?'  Simple fall:  'f(a\|e)ll(s\|ing\|en)?' |
| Original exclusion regex | ['fallopian', '(tube\|line\|catheter) f(a\|e)ll(s\|en\|ing\|ings)? out', '(level\|rate) f(a\|e)ll(s\|en\|ing\|ings)?', 'f(a\|e)ll(s\|en\|ing\|ings)? (\S+ )?(rate\|level)', '(level\|history of\|by a\|possible\|may have) (\S+ )?f(a\|e)ll(s\|en\|ing\|ings)?', 'f(a\|e)ll(s\|en\|ing\|ings)? (sustained \|last )?(\S+)?(month\|year\|jan(uary)? \|feb(uary)?\|mar(ch)?\|apr(il)?\|jun(e)?\|jul(y)?\|aug(ust)?\|sep(tember)?\|oct(ober)?\|nov(ember)?\|dec(ember)?)(s)?( ago)?', 'f(a\|e)ll(s\|en\|ing\|ings)? (from\|off\|off of) (\S+ )?(motor\|bi)cycle', 'f(a\|e)ll(s\|en\|ing\|ings)? asleep', 'f(a\|e)ll(s\|en\|ing\|ings)? clinic', 'frequent f(a\|e)ll(s\|en\|ing\|ings)?', 'f(a\|e)ll(s\|ing\|en\|ings)? level', 'not sure (that\|whether\|if) (\S+ )?f(a\|e)ll(s\|ing\|en\|ings)?', 'fall (and )?(return )?precaution(s)?', '(almost\|nearly\|would have) f(a\|e)ll(s\|ing\|en)?', 'faint (and\|&) fall', 'last fall', 'fall of', '(pressure\|map \|systolic) f(a\|e)ll(s\|ing\|en)?', 'f(a\|e)ll(s\|ing\|en)? (pressure\|map(s)? \|systolic)', '(AC\|anticoagulation\|lovenox\|NOAC\|warfarin\|coumadin\|rivaroxaban\|heparin\|eliquis\|dabigatran\|plavix\|aggrenox\|aspirin) (due to\|because of)( increased\| frequent \| risk of \| chances of \| chance of )? f(a\|e)ll(s\|en\|ing\|ings)?( risk)?', '(will\|would\|risk of\|chance of\|chances of) f(a\|e)ll(s\|en\|ing\|ings)?', 'f(a\|e)ll(s\|en\|ing\|ings)? off the wagon'] |
| New exclusion regex | ['f(a\|e)ll(s\|en\|ing)? away', 'f(a\|e)ll(s\|en\|ing)? out ', 'slips, trips, and falls', '200,000 of those falls', 'arter', 'history of', 'chance(s)?', 'pressure', 'farewell', 'fear of fall', 'fearful of fall', 'could have fallen', 'could fall', 'f(a\|e)ll(s\|en\|ing)? below', 'fall prec', 'predisposition', 'absence of', 'fall score', 'fall sign', 'assessment', 'next fall', 'afraid of fall', 'fall safety', 'previous fall(s)?', 'earlier this', 'precaution', 'agreement', 'video', 'teaching', 'prevent', 'risk', 'this fall', 'f(a\|e)ll(s\|en\|ing\|ings)? off', 'f(a\|e)ll(s\|en\|ing\|ings)? into', 'infallibilities', 'crestfallenness', 'fallaciousness', 'infallibility', 'crestfallenly', 'fallibilities', 'fallownesses', 'fallaciously', 'crestfallen', 'fallibility', 'fallaleries', 'infallible', 'fallacious', 'chapfallen', 'chopfallen', 'downfallen', 'chainfalls', 'fallboards', 'fallfishes', 'infallibly', 'nightfalls', 'fallowness', 'waterfalls', 'shortfalls', 'shortfall', 'waterfall', 'nightfall', 'fallboard', 'chainfall', 'befalling', 'evenfalls', 'downfalls', 'deadfalls', 'fallbacks', 'fallaways', 'fallalery', 'fallacies', 'infalling', 'footfalls', 'fallowing', 'windfalls', 'snowfalls', 'rockfalls', 'landfalls', 'rainfalls', 'pratfalls', 'rainfall', 'downfall', 'windfall', 'landfall', 'snowfall', 'fallible', 'fallback', 'pratfall', 'footfall', 'rockfall', 'fallaway', 'farfalle', 'deadfall', 'evenfall', 'fallfish', 'dewfalls', 'catfalls', 'befallen', 'icefalls', 'fallibly', 'falloffs', 'fallouts', 'fallowed', 'pitfalls', 'outfalls', 'unfallen', 'fallout', 'fallacy', 'pitfall', 'falloff', 'outfall', 'icefall', 'dewfall', 'befalls', 'catfall', 'fallals', 'fallers', 'infalls', 'fallows', 'fallow', 'befall', 'faller', 'infall', 'fallal', 'fellmongerings', 'fellowshipping', 'fellmongeries', 'fellmongering', 'schoolfellows', 'fellowshiping', 'fellowshipped', 'schoolfellow', 'fellowshiped', 'fellmongered', 'fellatrices', 'fellatrixes', 'fellmongers', 'fellmongery', 'fellowships', 'yokefellows', 'playfellows', 'fellowship', 'playfellow', 'yokefellow', 'fellnesses', 'bedfellows', 'fellmonger', 'fellations', 'bedfellow', 'fellating', 'fellaheen', 'fellatios', 'fellators', 'fellation', 'fellatrix', 'fellowing', 'fellowman', 'fellowmen', 'refelling', 'woolfells', 'fellatio', 'fellator', 'fellable', 'fellahin', 'fellated', 'fellates', 'fellowly', 'fellness', 'fellowed', 'woolfell', 'refelled', 'fellate', 'fellahs', 'felling', 'fellies', 'fellers', 'fellest', 'felloes', 'fellows', 'fellow', 'feller', 'fellah', 'felloe', 'felled', 'refell', 'fellas', 'befell', 'fella', 'felly', 'fells'] |
| Negations | ['no ', 'not ', "n't", 'negative', 'never', 'denies', 'deny'] |

### Appendix 2. Prompt examples

Figure 1. Zero-shot prompting example
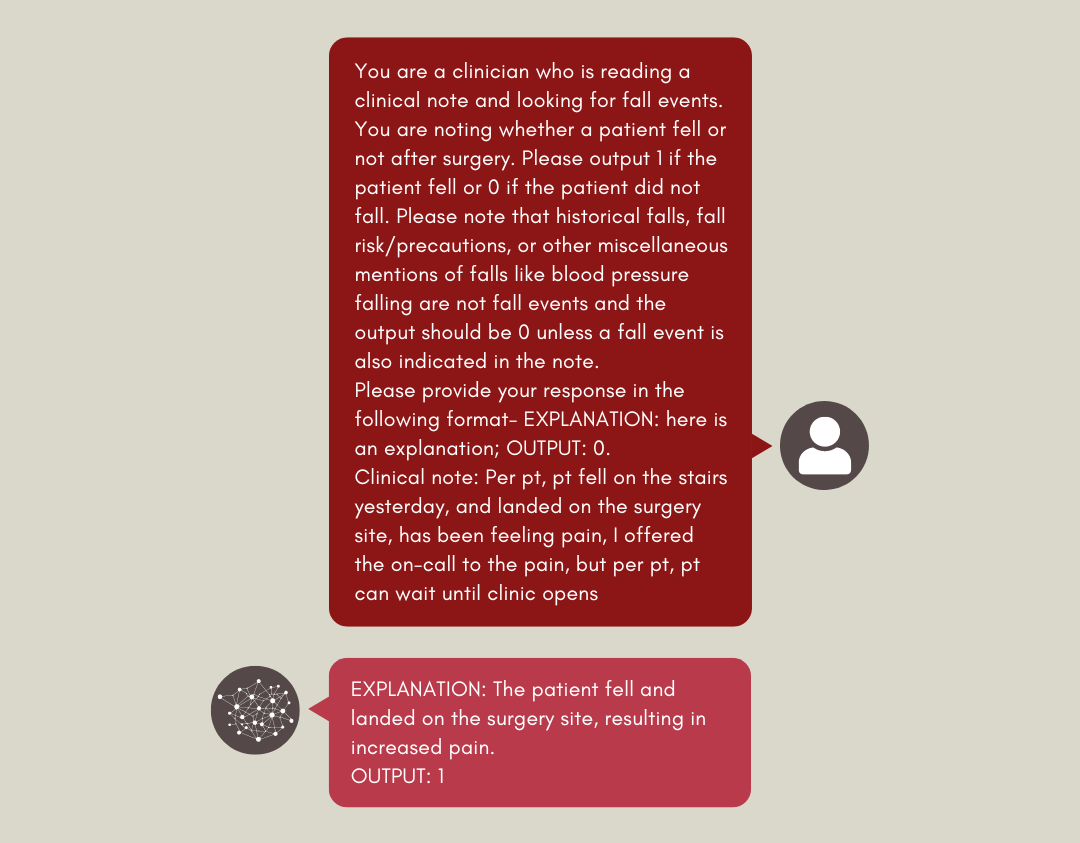

Figure 2. Few-shot prompting example

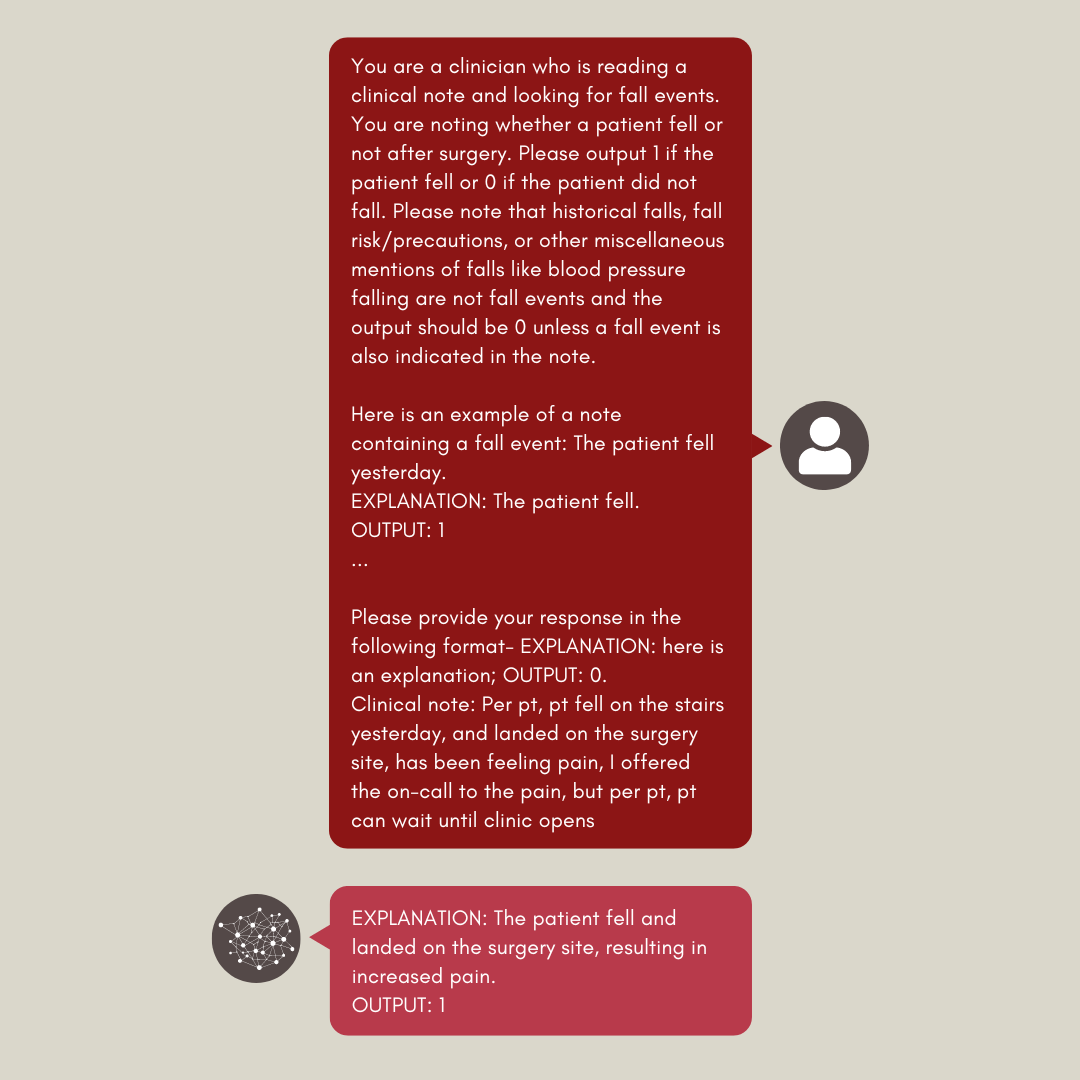
